# Somatic mutation involving diverse genes leads to a spectrum of focal cortical malformations

**DOI:** 10.1101/2021.12.22.21267563

**Authors:** Dulcie Lai, Meethila Gade, Edward Yang, Hyun Yong Koh, Nicole M. Walley, Anne F. Buckley, Tristan T. Sands, Cigdem I. Akman, Mohamad A. Mikati, Guy M. McKhann, James E. Goldman, Peter D. Canoll, Allyson L. Alexander, Kristen L. Park, Gretchen K. Von Allmen, Meenakshi B. Bhattacharjee, Hart G.W. Lidov, Hannes Vogel, Gerald A. Grant, Brenda E. Porter, Annapurna H. Poduri, Peter B. Crino, Erin L. Heinzen

## Abstract

Post-zygotically acquired genetic variants, or somatic variants, that arise during cortical development have emerged as important causes of focal epilepsies, particularly those due to malformations of cortical development. Pathogenic somatic variants have been identified in many genes within the PI3K-AKT3-mTOR-signaling pathway in individuals with hemimegalencephaly and focal cortical dysplasia (type II), and more recently in *SLC35A2* in individuals with focal cortical dysplasia (type I) or non-dysplastic epileptic cortex. Given the expanding role of somatic variants across different brain malformations, we sought to delineate the landscape of somatic variants in a large cohort of patients who underwent epilepsy surgery with hemimegalencephaly or focal cortical dysplasia. We evaluated samples from 123 children with hemimegalencephaly (n=16), focal cortical dysplasia type I and related phenotypes (n=48), focal cortical dysplasia type II (n=44), or focal cortical dysplasia type III (n=15) classified using imaging and pathological findings. We performed high-depth exome sequencing in brain tissue-derived DNA from each case and identified somatic single nucleotide, indel, and large copy number variants. In 75% of individuals with hemimegalencephaly and 29% with focal cortical dysplasia type II, we identified pathogenic variants in PI3K-AKT-mTOR pathway genes. Four of 48 cases with focal cortical dysplasia type I (8%) had a likely pathogenic variant in *SLC35A2*. While no other gene had multiple disease-causing somatic variants across the focal cortical dysplasia type I cohort, four individuals in this group had a single pathogenic or likely pathogenic somatic variant in *CASK, KRAS, NF1*, and *NIPBL*, genes associated with neurodevelopmental disorders. No rare pathogenic or likely pathogenic somatic variants in any neurological disease genes like those identified in the focal cortical dysplasia type I cohort were found in 63 neurologically normal controls (*P* = 0.017), suggesting a role for these novel variants. We also identified a somatic loss-of-function variant in the known epilepsy gene, *PCDH19*, present in a very small number of alleles in the dysplastic tissue from a female patient with focal cortical dysplasia IIIa with hippocampal sclerosis. In contrast to focal cortical dysplasia type II, neither focal cortical dysplasia type I nor III had somatic variants in genes that converge on a unifying biological pathway, suggesting greater genetic heterogeneity compared to type II. Importantly, we demonstrate that FCD types I, II, and III, are associated with somatic gene variants across a broad range of genes, many associated with epilepsy in clinical syndromes caused by germline variants, as well as including some not previously associated with radiographically evident cortical brain malformations.

## Introduction

The cerebral cortex is assembled during embryonic development through a series of tightly regulated processes that include neuronal and glial progenitor cell proliferation, cellular differentiation and fate specification, neuronal migration, and ultimately cortical organization. Disruption of any of these processes may result in a range of cortical malformations, spanning those that affect all or most of the cortex to small focal cortical lesions that may or may not be detectable with brain imaging (Barkovich *et al*., 1996; Barkovich *et al*., 2012). Regardless of the size of the lesion, malformations of cortical development (MCD) are commonly associated with refractory epilepsy (Leventer *et al*., 1999), as well as intellectual disability and autism spectrum disorder (Casanova *et al*., 2013).

Most MCD are thought to arise from variants in genes that encode proteins essential for neurodevelopment. Pathogenic genetic variants that result in MCD can be inherited, newly acquired in the germ cell (gamete) of a parent and transmitted to the affected child and thus appearing as *de novo* variants not detected in the parents’ blood- or buccal-derived DNA, or post-zygotically acquired during embryonic development, giving rise to a mosaic pattern of variant-positive and variant-negative cells and appearing as somatic variants with variant allelic fractions (VAFs) less than 50%. Typically, *de novo* germline variants or very early (pre-gastrulation) post-zygotically acquired somatic variants lead to more diffuse cortical malformations (Guerrini and Dobyns, 2014; Jamuar *et al*., 2014; D’Gama *et al*., 2017), whereas somatic variants have been commonly identified in some forms of focal MCD (Lee *et al*., 2012; Poduri *et al*., 2012; Winawer *et al*., 2018).

Focal cortical dysplasia (FCD) is a type of MCD characterized by focal disruption of cortical cytoarchitecture that is highly associated with medication-resistant epilepsy. The International League Against Epilepsy has provided a classification system to define FCD lesions according to pathological findings in resected brain tissue specimens. Specifically, abnormalities in radial or tangential cortical lamination are deemed Type I (FCDI), the presence of dysmorphic neurons and/or balloon cells indicate Type II (FCDII), and cortical lamination abnormalities in combination with another brain lesion indicate Type III (FCDIII) (Palmini *et al*., 2004; Blumcke *et al*., 2011). Recently, a new pathological entity has been described as a mild malformation of cortical development (mMCD) in the presence of oligodendroglial hyperplasia (MOGHE), as evidenced by excess Olig2 and PDGFRα-immunoreactive oligodendroglia and heterotopic white matter neurons. The histological features of this newly defined pathology in resected tissue specimens may be subtle and can range from FCDI to mMCD to gliosis only (Schurr *et al*., 2017). It should be noted that clinically, pre-operative MRI is sufficient to make the diagnosis of hemimegalencephaly (HMEG) and frequently to make a presumptive diagnosis of FCD, with FCDII frequently associated with a ‘transmantle’ sign consisting of abnormal signal extending from the ventricular region to the cortex. Post-operatively, a more definitive diagnosis can be made that incorporates available pathological findings, which may thus result in reclassification from the initial diagnosis. This is particularly relevant for ‘nonlesional’ (MRI-negative) cases that are then found to have pathological evidence of FCD.

Somatic variants in several genes in the PI3K-AKT-mTOR signaling pathway have been identified in individuals with FCDII and hemimegalencephaly (Lee *et al*., 2012; Poduri *et al*., 2012; Lim *et al*., 2015; Nakashima *et al*., 2015; Moller *et al*., 2016; Lim *et al*., 2017; Pelorosso *et al*., 2019; Salinas *et al*., 2019; Zhao *et al*., 2019), and have been reported in one study to account for approximately 90% of hemimegalencephaly and 60% of FCDII cases (Baldassari *et al*., 2019). Recently somatic variants in *SLC35A2* have been identified in resected brain tissue found to have pathology consistent with FCDI and mMCD, but also in individuals with histologically normal tissue (Sim *et al*., 2018; Winawer *et al*., 2018; Bonduelle *et al*., 2021). The reasons for the variable pathologic phenotypes across studies is unclear but may reflect variability in the extent of mosaicism in tissue or within specific cell types (Winawer *et al*., 2018; Heinzen, 2020; Miller *et al*., 2020; Bonduelle *et al*., 2021), as well as the challenges with accurately assigning histopathological diagnoses across subtle and highly localized phenotypes characteristic of FCDI and mMCD pathologies (Chamberlain *et al*., 2009). To date, mosaic loss-of-function variants in *SLC35A2* explain between 17-29% of cases of refractory neocortical epilepsy with and without FCDI pathology (Sim *et al*., 2018; Winawer *et al*., 2018; Bonduelle *et al*., 2021). *SLC35A2* encodes what is believed to be the sole transporter of UDP-galactose from the cytosol to the Golgi apparatus, where it can be acted on by galactosyl-transferases to generate galactose-containing glycoproteins, proteoglycans, and glycolipids. Absence of this transporter is expected to have wide-reaching effects on protein and lipid glycosylation. Before the discovery of somatic *SLC35A2* variants in focal neocortical epilepsy, germline *SLC35A2* variants had been associated with severe epileptic encephalopathies and intractable seizures (Kodera *et al*., 2013; Ng *et al*., 2013).

Here we report the results of a genome-wide analysis for somatic single nucleotide variants (SNV), small insertion or deletion variants (indels) in protein-coding regions, and large copy number variants (CNVs) in resected brain tissue samples from 123 individuals with epilepsy who underwent focal surgical resection for medication-resistant epilepsy and genetic sequencing of the resected tissue. Our study demonstrates that at least a portion of FCDI and related MCD are due to somatic, brain-tissue-specific genetic variants in genes previously implicated in epilepsy and other neurodevelopmental disorders, with the associated focal epilepsy phenotypes likely reflecting the tissue and cellular localization of the pathogenic genetic variants identified. Notably, the types of gene variants involved in FCDI and FCDIII were distinct from those characteristic of FCDII providing a new molecular genetic understanding of FCD pathogenesis.

## Materials and methods

### Study participants

#### MCD cohort

Individuals who underwent a surgical resection of cortical tissue due to intractable focal epilepsy were enrolled from Boston Children’s Hospital, Duke University Medical Center, Lucille Packard Children’s Hospital at Stanford, The University of Texas Health Science Center at Houston, or Children’s Hospital Colorado. Clinical assessment of the candidacy for focal resection was determined by standard clinical practice at each site, incorporating seizure semiology, EMG/EEG data, structural MRI, functional imaging (PET, SPECT), and consensus at epilepsy surgery conferences.

To be included in this study, patients were required to have no known genetic syndrome at the time of enrollment and have a radiographic phenotype that was not inconsistent with MCD, which includes cases without evident focal lesions. Exclusion criteria included encephalomalacia, Rasmussen’s encephalitis, isolated schizencephaly, neoplasm, isolated mesial temporal lobe sclerosis, gray matter heterotopia, polymicrogyria, and clinically diagnosed tuberous sclerosis complex. Imaging and neuropathological data or reports were reviewed by a neuroradiologist (EY), neuropathologist (HGWL), and epileptologist/neurogeneticist (AP) to determine eligibility for this study and classify each eligible case into one of four phenotypic categories: first, FCDI^+^, represents cases with focal epilepsy with MRI evidence of FCD with or without available neuropathological features of FCDI. Included in the FCDI+ group were cases with MRI lesions suggestive of FCD type I and neuropathological evidence of FCD type I and related subtypes, e.g. FCD Ia, Ib, and Ic, mMCD. Additionally cases deemed FCD not otherwise specified (FCDNOS), with MRI lesions suggestive of FCD type I but non-specific histopathology, e.g. gliosis, or no pathological abnormalities and cases with non-lesional focal epilepsy (NLFE), defined as normal brain MRI, with FCD type I or related pathology *or* without FCD type I or related pathology (maintaining the assumption that the pathology was likely present but not observed due to incomplete sampling of the specimen), were included. The second category was FCDII and included cases with FCD types IIa and IIb, with or without FCD detected on MRI. The third category was FCDIII and included cases with FCDIIIa, FCDIIIc and FCDIIId. The fourth was HMEG, including cases with classical HMEG as well as cases with dyplastic megalencephaly (DMEG) in the form of large malformations that do not fully encompass an entire hemisphere or that also involve both cerebral hemispheres. The clinical diagnoses of all individuals are provided in Supplementary Table 1. All patients were consented to participate in research approved by the respective institutional review boards. Specifically, the Duke University Health System Institutional Review Board, the Stanford University Institional Review Board, the Boston Children’s Hospital Institutional Review Board, the University of Texas Health Science Center at Houston Institional Review Board, and the Columbia University Irving Medical Center Institutional Review Board all reviewed and approved the research studies, including specifics of how individuals would be consented, how patient samples would be used, how privacy is maintained, and how data generated from their specimens and clinical data will be used. A subset of the patients have been reported in previous publications (Supplementary Table 1).

#### Control cohort

We obtained sequencing data from brain tissue specimens obtained from autopsies from controls without neurological disorders. Three-hundred forty-two exome-sequenced individuals were obtained from the North American Brain Expression Consortium (NABEC - dbGaP Study Accession: phs001300.v1.p1), and 15 whole genome-sequenced controls were downloaded from the Brain Somatic Mosaicism Network (National Institute of Mental Health Data Archive) (Supplementary Table 2).

#### Specimen collection and DNA extraction

Brain tissue specimens were collected from each of the individuals in the MCD cohort. Blood samples were collected when possible (Supplementary Table 3). DNA was extracted from the specimens using GenFind V3 (Beckman Coulter) or DNeasy Blood & Tissue Kits (Qiagen) per the manufacturer’s protocol.

### Sequencing and variant calling

#### Exome sequencing

Brain-tissue-derived DNA samples from the 123 individuals underwent paired-end high-depth exome sequencing on an Illumina HiSeq2000, HiSeq2500, or NovaSeq at Duke University Medical Center (DUMC), Columbia University Irving Medical Center (CUIMC), Genewiz, or the University of North Carolina at Chapel Hill (UNC) high throughput sequencing facilities. The protein-coding sequences were enriched in the libraries using either Nimblegen SeqCap EZ V3.0, Agilent V6 SureSelect, IDT xGen Exome Research Panel v1, or IDT xGen Exome Research Panel v2 exome enrichment kits. We also performed exome sequencing on DNA extracted from whole blood samples for 74 individuals (Supplementary Table 3).

#### Alignment

Next-generation sequence data from cases and controls were aligned to the hg38 reference genome (GRCh38.d1.vd1) using Burrows-Wheeler Aligner (BWA, version 0.7.15) (Li and Durbin, 2009). Biobambam2 (version2.0.168) and samtools (1.11) (Li *et al*., 2009) were used for marking duplicates, sorting, and indexing alignment files.

#### Variant calling

##### Somatic single-nucleotide and small insertion-deletion variant calling

SNV and indel variants were called in the brain-derived DNA samples using Mutect2 (Cibulskis *et al*., 2013) following the NCI Genomics Data Commons Somatic Variant Calling Workflow. The pipeline included an assessment of a panel of normal and germline variants compiled from the gnomAD database (gnomad.hg38.vcf.gz) (Karczewski *et al*., 2020) to reduce artifactual calls. While matched blood was exome sequenced in some cases, only exome sequencing performed in brain DNA was used in the primary analysis to ensure uniform analysis of all study subjects. Since the algorithms used variant calling in the absence and presence of a matched leukocyte-derived DNA sample, when exome sequence data from a matched leukocyte-derived DNA sample was available, brain tissue somatic variants were also called in the pair and compared to the results from the brain-only calling to ensure that candidate variants were not overlooked.

High-quality somatic SNVs and indels were defined as the subset of somatic variant calls meeting the following criteria: (1) annotated to be located in a protein-coding exon or associated splice sites; (2) flagged as “PASS” by the Mutect2 software; (3) called at a site with at least 20 reads covering the site; (4) at least five reads supporting the variant allele; (5) reads supporting the variant were found on both read 1 and read 2 of the paired-end sequencing of a DNA fragment; (6) variant allele fraction (VAF < 35%) for calls on autosomes and the X chromosome of females, and VAF < 70% for chromosome X variants in a male; and a (7) VAF > 2%. Variants on the Y chromosome and those where multiple different variant alleles were called in the same individual were also excluded.

Somatic SNVs and indels that did not meet the strict high-quality criteria (“permissive” variant calls) were also evaluated for candidate genetic diagnoses. These permissive calls included all somatic variants called by the Mutect2 algorithm in a protein-coding exon or splice site, excluding only those suggestive of a sequencing or alignment error (*i*.*e*. variants failing the base_qual, clustered_events, contamination, map_qual, or strand_bias filters, or variants called as multi-allelic).

Somatic SNV and indel variant calls were inspected for accurate calling using Integrative Genomics Viewer (IGV, version 2.8.7).

##### Somatic copy number variant calling

Somatic copy number variants were called in the brain tissue-derived DNA using CNV Radar (Cibulskis *et al*., 2013). This algorithm uses a panel of normal samples, read depth, and variant allele fraction (VAF) patterns to detect CNVs in the absence of matched controls. Called CNVs were annotated using ANNOT-SV (Geoffroy *et al*., 2018). Given the challenges of accurately detecting somatic CNVs from exome sequencing data, we limited these analyses only to large >1 Mb CNVs.

##### Germline single nucleotide and small insertion-deletion variant calling

Germline indels and SNVs were called using GATK Best Practices recommendations (DePristo *et al*., 2011; Van der Auwera *et al*., 2013). The process included base quality scores recalibration to generate analysis ready reads, individual level variant calling using HaplotypeCaller, indel realignment, duplicate removal, and joint genotyping across the full cohort. The resultant SNV and indel calls were then hard filtered using QD < 2.0 || FS > 60.0 || MQ < 40.0 || MQRankSum < -12.5 || ReadPosRankSum < -8.0,, and QD < 2.0 || FS > 200.0 || ReadPosRankSum < -20.0, respectively.

#### Quality control of exome sequence data

Somalier (version 0.2.12) was used to infer the sex and ancestry of each sequenced individual and to test for unexpected relatedness among cases and controls (Pedersen *et al*., 2020). Ancestry was predicted on the query samples using the dataset from the 1000 genomes project-hg38 as a reference (Genomes Project *et al*., 2015). We also assessed the rate of G-T substitutions which are a well-recognized Illumina sequencing error and can also arise from oxidative DNA damage in low-quality DNA samples (Costello *et al*., 2013; Chen *et al*., 2017).

Average coverage and percent of bases covered at least 50-fold across the CCDS protein-coding regions plus two base pairs flanking exons were calculated for each sample using the GATKDepthOfCoverage tool including the GATK analysis toolkit.

#### Annotation

Annotation of the somatic and germline variants was performed using ANNOVAR (v20200609) (Wang *et al*., 2010). Annotation databases used were either downloaded directly from ANNOVAR or created using the original database (Supplementary Table 4).

Protein-coding variants analyzed in this study were limited to only those located in the Consensus Coding Sequence (CCDS version 22, hg38) (Pruitt *et al*., 2009) and two base pair splice site regions flanking the CCDS exons.

#### Variant confirmation

Somatic variants were confirmed using digital PCR (dPCR), Sanger sequencing, or amplicon sequencing. Digital PCR was performed using custom-designed Taqman assays targeting the variant and reference alleles for an SNV variant or targeting a gene within the duplicated or deleted region for a CNV. A PCR QuantStudio 3D Digital PCR System (Thermo Fisher Scientific) was used to quantitatively assess allele fraction and copy number per the manufacturer’s protocol.

In some cases where the candidate somatic variant had a high VAF estimated from exome sequencing or a Taqman assay could not be designed, we confirmed variants with either: (1) PCR followed by Sanger sequencing; (2) allele-specific PCR whereby two forward primers were designed to each selectively amplify the variant or the reference sequence in the presence of the same reverse primer placed downstream of the variant; or (3) amplicon sequencing (CDG Genomics or Genewiz) which entailed PCR amplification of a >100 bp fragment followed by targeted next-generation sequencing using an Illumina MiSeq to ∼1000-fold coverage of the variant site and flanking genomic regions. All PCR reactions used primers designed using Primer3 and purchased from Integrated DNA Technologies. MyTaq Mix (Meridian Bioscience) was used in all PCR reactions according to the manufacturers’ instructions. Sanger sequencing was performed by Genewiz. In one individual (neuro1410F49br), the somatic *MTOR* variant identified in the brain tissue samples was confirmed clinically with amplicon sequencing in an independent sample.

### Diagnostic analyses

#### Somatic variant diagnoses

We evaluated the following sets of variant calls to comprehensively search for candidate somatic genetic diagnoses:

1. high-quality somatic single-nucleotide and indel variants (defined above) that were rare [gnomAD exome minor allele frequency (MAF) <1 × 10^−6^], putatively functional [splice-site, nonsense, missense excluded Polyphen (HumVar) benign, frameshift and nonframeshift indel], and located in an Online Mendelian Inheritance in Man (OMIM) dominant neurological disease gene, defined as any gene associated with disorders for which a neurologic phenotype entry was included in the clinical synopsis and excluding genes associated with autosomal recessive disorders (Supplementary Table 5)
2. permissive somatic single nucleotide substitution and indel variants (defined above) that were classified as pathogenic or likely pathogenic in ClinVar located in an OMIM neurological disease gene and all loss-of-function variants (splicing, nonsense, or frameshift indel) in genes classified as haploinsufficient in the ClinGen Dosage Sensitivity Map (https://dosage.clinicalgenome.org) (Supplementary Table 5)
3. permissive somatic single nucleotide substitution and indel variants that are rare [gnomAD exome MAF <1 × 10^−6^] and putatively functional [splice-site, nonsense, missense excluded Polyphen (HumVar) benign, frameshift and nonframeshift indel] in known MCD genes (*AKT3, CASK, DEPDC5, KRAS, MTOR, NF1, NIPBL, PCDH19, PIK3CA, PTEN, RHEB, SLC35A2, TSC1*, and *TSC2*)
4. somatic CNV calls >1 Mb

We assessed the likelihood that each variant meeting the above criteria contributed to the corresponding patient’s clinical phenotype. The genotype of those deemed as possible genetic diagnoses was then independently confirmed as a true somatic variant using an orthogonal genotyping approach (see above). Variants were classified according to recently defined ClinVar variant curation rules that govern the pathogenicity assessment of somatic variants specifically related to developmental brain malformations (ClinGen Brain Malformation Expert Panel).

#### Germline variant diagnoses

While the primary focus of this study was to identify disease-causing somatic variants in MCD, we also evaluated the sequence data for definitive germline diagnoses since there are germline genetic causes of some MCD and for comparison to the somatic variant analysis. Among the high-quality germline variant calls, we searched for pathogenic or likely pathogenic variants that met American College of Medical Genetics (ACMG) guidelines (Richards *et al*., 2015), including all variants classified as pathogenic or likely pathogenic in ClinVar and all loss-of-function variants (splicing, nonsense, or frameshift indel) in genes classified as haploinsufficient in the ClinGen Dosage Sensitivity Map. Each variant was then assessed the likelihood that those variants contribute to the patient’s phenotype.

### Gene set enrichment analyses

#### Gene sets analyzed

The burden of variants in all cases compared to controls was assessed across multiple gene sets (Supplementary Table 5), including: (1) Online Mendelian Inheritance in Man (OMIM) dominant neurological disease-associated genes as defined above; (2) OMIM non-neurological disease genes, including all OMIM morbid genes and excluding any that have a neurologic phenotype entry included in the clinical synopsis; (3) loss-of-function intolerant genes (pLI>0.9 - gnomAD database (v2.0.1)); (4) constrained genes (missense Z score >3.09, gnomAD database (v2.0.1)); and (5) Kyoto Encyclopedia of Genes and Genomes (KEGG) neurological pathways (Kanehisa and Goto, 2000).

#### Coverage normalization across cases and controls

Before performing burden testing in the case and control cohorts, sample- and locus-level coverage harmonization was performed to minimize the systemic bias due to differences in sequencing depths between the case and control cohorts. First, subjects were excluded if <80% of the protein-coding regions were sequenced at least 50-fold. This exclusion left us with 63 controls and 113 MCD cases [FCDI^+^ (n=46), FCDII (n=39); FCDIII (n=13), and HMEG (n=15)]. To further address the imbalance in coverage between cases and controls, individual site-level coverage levels were generated using the GATKDepthOfCoverage tool at each site in the exonic and 2-bp splice site regions included in the CCDS. Within the case and control cohorts separately, we identified the sites in the autosomes with at least 50 reads in 90% of the samples. To incorporate the differences in depth of sequencing on the X chromosome in males and females, sites were excluded if they did not have at least 25 reads in males and 50 reads in females in 90% of the cohort. After the case and control cohorts were separately normalized, we then selected only sites that were covered per the defined criteria across 90% of the combined case and control cohort. Finally, the coverage normalized sites were then collapsed by CCDS transcripts and genes, and only genes with at least 50% normalized coverage were included in the analysis (n=15,622, Supplementary Table 6).

#### Gene set enrichment testing

Gene set enrichment analyses were performed on coverage normalized calls using DNENRICH (Fromer *et al*., 2014). The software models exome-wide dispersion of high-quality somatic variants under a null hypothesis that accounts for gene size, the tri-nucleotide context of single nucleotide substitutions, and the functional effect of mutations.

In brief, somatic variants identified in each individual were randomly distributed among the coverage normalized gene set based on the trinucleotide sequence and the functional effect of a mutation (i.e. a single base substitution ATC to AGC that results in a missense variant could be placed at any ATC site located in any of the coverage normalized genes that when the T is substituted for a G would result in a missense variant). The somatic variants identified in each individual were randomly placed in the exome in this way 10,000 times and a one-sided statistical test was performed to compare the observed data to the simulated datasets within a cohort. The relative proportion of qualifying mutations in a gene set in cases was then compared to that of controls. This analysis was performed in the FCDI^+^, FCDII, FCDIII, and HMEG cohorts. We tested for enrichment of somatic variants within sets of genes including KEGG pathway genes, constrained genes, LOF intolerant genes, OMIM dominant neurological genes, and OMIM non-neurological disease genes (Supplementary Table 5) compared to controls.

### Data availability

Exome sequence data from cases evaluated in this study have been deposited into dbGAP (submission pending).

## Results

### Case ascertainment and phenotypic description of the cohort

One hundred forty-seven patients with refractory focal epilepsy who had undergone resective epilepsy surgery were evaluated for inclusion in this study. An initial assessment of brain tissue sample quality led to five cases being excluded due to failed quality control checks after sequencing, including one gender mismatch, and four due to excessive rates of G-T substitutions indicative of poor-quality DNA.

Each of the 142 remaining cases underwent a centralized review of pre-surgical MRI and neuropathological findings. Nineteen cases were excluded because they did not meet inclusion criteria, e.g. MRI showed evidence of stroke or hypoxic-ischemic injury. The remaining cases comprised our cohort of 123 individuals. Cases were reviewed and classified by consensus a FCDI^+^ (n=48), FCDII (n=44), and FCDIII (n=15), or HMEG (n=16) (Figure 1A, Supplementary Table 1). A summary of the clinical phenotypes for each of these 123 individuals is provided in Supplementary Table 1. Representative MRIs are shown in Figure 2.

**Figure 1.**
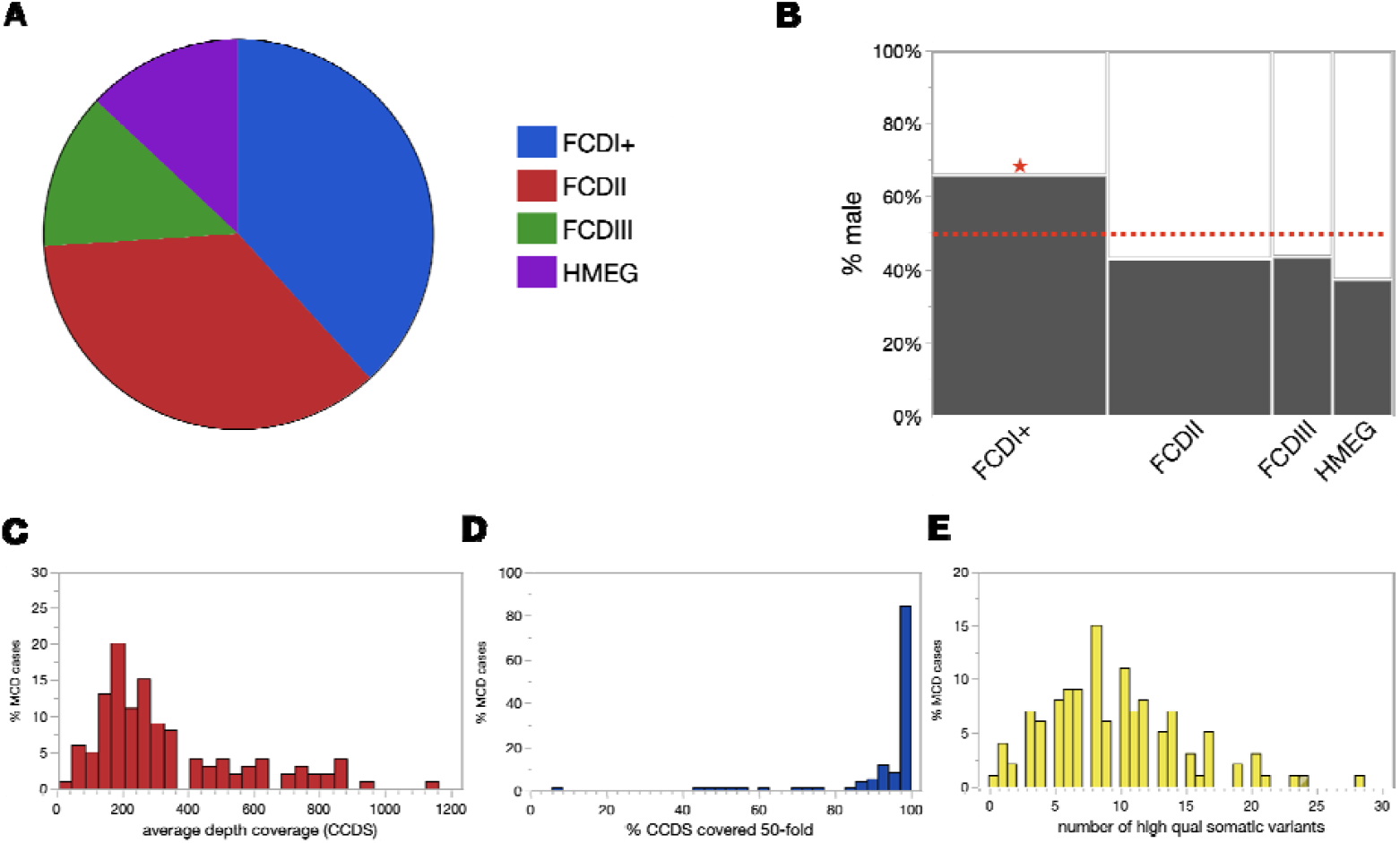
Summary of MCD cohort. (A) Percentages of patients binned in broadly-defined MCD phenotypes. (B) Fraction of males within each MCD phenotype (size of the bar corresponds to size of the cohort). (C) Average depth of coverage, percent of protein-coding regions sequenced at least 50-fold, and number of high quality somatic variants called in each subject.

**Figure 2.**
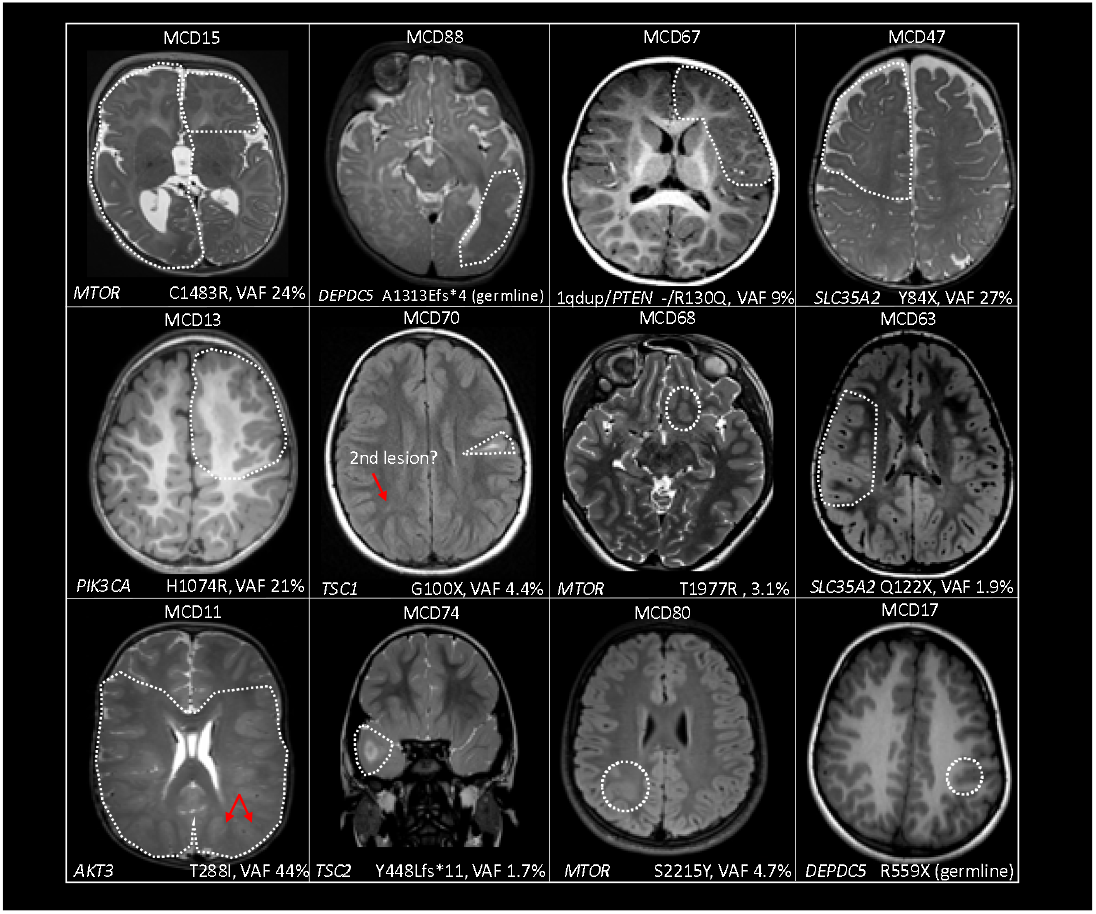
Representative MRI images from MCD cases.

A subset of FCD specimens (n=12) included tissue resected from individuals with no discernable lesion on pre-operative MRI who were thus pre-operatively delineated as NLFE. Pathological review of these NLFE cases revealed findings consistent with FCDI or, mMCD (n=5), FCDII (n=2), or FCDIIIc (n=1), while four individuals had no clear neuropathological abnormalities. The five FCDI/mMCD cases and the pathologically normal subset (n=4) were included in the FCDI^+^ category. The three cases with pathology consistent FCDII and III were binned in the FCDII and FCDIII cohorts, respectively, despite being radiographically non-lesional (Supplementary Table 1).

There were approximately equal numbers of males (n=63) and females (n=60) across the whole cohort, but we observed more males in the FCDI^+^ category (66%, *P* = 0.011) and there was a trend toward more females in the FCDII and HMEG cohorts, (Figure 1B).

### Exome sequencing technical summary

The brain tissue samples from the above 123 individuals were sequenced to an average depth of 367-fold; 122 (99%) had 80% of protein-coding regions covered at least 50-fold. Each sample had on average nine high-quality somatic SNVs or indel variants in the protein-coding regions of the genome (Figure 1C). Among the high-quality somatic variants that were selected for confirmation, we validated 24/26 (92%) using an orthogonal genotyping approach (Supplementary Table 7).

### Targeted somatic diagnostic analysis

Given that we expected a fraction of the MCD cases to harbor a known pathogenic variant in either *SLC35A2* or a PI3K-AKT-mTOR signaling pathway gene, we first performed a targeted ‘diagnostic’ analysis to identify pathogenic or likely pathogenic variants based on American College of Medical Genetics (ACMG) guidelines (Richards *et al*., 2015).

#### HMEG

Somatic gene variants were detected in 12 of 16 (75%) HMEG cases (Table 1). Six individuals each had a somatic *PIK3CA* variant, including five resulting in the previously reported recurrent E545K amino acid substitution and one with the previously reported recurrent H1047R substitution (Lee *et al*., 2012; D’Gama *et al*., 2015). Three individuals each had a recurrent somatic *MTOR* variant, two resulting in C1483R and one in C1483Y at the protein level (Lee *et al*., 2012). Two cases each had a somatic variant in *AKT3*, one resulting in the previously reported E17K variant (Lee *et al*., 2012; Poduri *et al*., 2012) and one with a novel variant resulting in *AKT3* T288I. All except the T288I *AKT3* variant were identified as high-quality variant calls. The *AKT3* T288I variant (VAF = 42%) only in the paired brain-blood variant calls because it was presumed germline in the brain-only call set given the relatively high VAF. The *AKT3* T288I variant was absent in the leukocyte-derived DNA exome sequencing data, with 300-fold coverage at the variant site, and in data obtained by digital PCR. Despite the lack of detectable variant in the blood, the patient presented clinically with neurological and non-neurological features of a segmental overgrowth syndrome including a capillary hemangioma and toe syndactyly (Table 2), suggesting that the variant may have been present outside of the central nervous system.

**Table 1.** List of pathogenic and candidate disease-causing somatic variants identified.

| Sample | Sex | Phenotype category* | Gene - Variant | ACMG classification (evidence) | Somatic/germline | Brain DNA VAF (exome) [95% CI] | Brain DNA VAF (dPCR/amplicon seq) [95% CI] | Blood DNA VAF (exome), [95% CI] |
| --- | --- | --- | --- | --- | --- | --- | --- | --- |
| MCD5 | F | HMEG | 16pUPD | LP | somatic | - | - | - |
| MCD11 | M | HMEG | *AKT3* - NM_181690 - c.C863T;p.T288I | P (PS2, PM1, PM2, PP2, PP3) | somatic | 44% [38 - 50%] | - | 0% [0 - 1.2%] |
| MCD6 | F | HMEG | *AKT3* - NM_181690 - c.G49A;p.E17K | P (PS2, PM1, PM2, PP2, PP3, PP5) | somatic | 13% [9.8 - 19%] | - | 0% [0 - 1.9%] |
| MCD9 | F | HMEG | *MTOR* - NM_004958 - c.G4448A;p.C1483Y | P (PS2, PM1, PM2, PM5, PP3, PP5, BP1) | somatic | 11% [8.2 - 15%] | 12% [11 - 13%] | - |
| MCD15 | M | HMEG | *MTOR* - NM_004958 - c.T4447C;p.C1483R | P (PS2, PM1, PM2, PM5, PP3, PP5, BP1) | somatic | 24% [20 - 28%] | 29% [27 - 31%] | 2% [0 - 4.7%] |
| MCD4 | F | HMEG | *MTOR* - NM_004958 - c.T4447C;p.C1483R | P (PS2, PM1, PM2, PM5, PP3, PP5, BP1) | somatic | 5.7% [1.9 - 13%] | - | 0% [0 - 6%] |
| MCD2 | F | HMEG | *PIK3CA* - NM_006218 - c.G1624A;p.E542K | P (PS2, PM1, PM2, PM5, PP2, PP3, PP5) | somatic | 14% [11 - 18%] | 19% [17 - 21%] | 0% [0 - 0.6%] |
| MCD10 | F | HMEG | *PIK3CA* - NM_006218 - c.G1624A;p.E542K | P (PS2, PM1, PM2, PM5, PP2, PP3, PP5) | somatic | 25% [17 - 36%] | 10% [9.2 - 11%] | - |
| MCD3 | F | HMEG | *PIK3CA* - NM_006218 - c.G1633A;p.E545K | P (PS2, PS3, PM1, PM2, PM5, PP2, PP3, PP5) | somatic | 28% [24 - 34%] | 23% [21 - 26%] | 0% [0 - 1.1%] |
| MCD12 | M | HMEG | *PIK3CA* - NM_006218 - c.G1633A;p.E545K | P (PS2, PS3, PM1, PM2, PM5, PP2, PP3, PP5) | somatic | 20% [15 - 27%] | 22% [20 - 24%] | 0% [0 - 2.5%] |
| MCD14 | M | HMEG | *PIK3CA* - NM_006218 - c.G1633A;p.E545K | P (PS2, PS3, PM1, PM2, PM5, PP2, PP3, PP5) | somatic | 14% [6.8 - 25%] | 27% [26 - 28%] | - |
| MCD13 | M | HMEG | *PIK3CA* - NM_006218 - c.A3140G;p.H1047R | P (PS2, PM1, PM2, PM5, PP2, PP3, PP5) | somatic | 21% [17 - 26%] | 14% [13 - 16%] | - |
| MCD39 | M | FCDI<sup>+</sup> | 1qdup | LP | somatic | - | - | - |
| MCD59 | M | FCDI<sup>+</sup> | *ATP10A* - NM_024490 - c.C647A;p.S216X | VOUS | somatic | 2.9% [1.6 - 5.0%] | 1.5% [1.2-1.9] | - |
| MCD20 | F | FCDI<sup>+</sup> | *CASK* - NM_001367721 - c.2317+1G>A | P (PVS1, PS2, PM2, PP3, PP5) | somatic | 6.5% [4.5 - 9.2%] | 4.8% [2.6 - 9.5%] | 0% [0 - 1.6%] |
| MCD40 | M | FCDI<sup>+</sup> | *CUL1* - NM_001370660 - c.C187T;p.R63X | VOUS | somatic | 7.2% [3.5 - 13%] | 5.5% [4.8 - 6.3%] | 0% [0 - 3.6%] |
| MCD17 | M | FCDI<sup>+</sup> | *DEPDC5* - NM_001369901:c.C1675T:p.R559X | P (PVS1, PS2, PM2, PP3, PP5) | germline | 50% [42 - 58%] | - | - |
| MCD60 | M | FCDI<sup>+</sup> | *KRAS* - NM_001369786 - c.G35T;p.G12V | P (PS2, PS3, PM1, PM2, PM5, PP2, PP3, PP5) | somatic | 20% [15 - 25%] | 18% [17 - 19%] | - |
| MCD54 | M | FCDI<sup>+</sup> | *NF1* - NM_000267 - c.1017_1018del;p.S340Cfs*12 | P (PVS1, PS2, PM2, PP3, PP5) | somatic | 3.0% [1.7 - 4.7%] | 2.1% [1.6 - 2.6%] | 0%[0-0.06%]** |
| MCD21 | F | FCDI<sup>+</sup> | *NIPBL* - NM_015384 - c.7685+1G>A | LP (PVS1, PM2, PS2) | somatic | 6.7% [2.5 - 14%] | 16% [15 - 18%] | 0% [0 - 2%] |
| MCD38 | M | FCDI<sup>+</sup> | *SLC35A2* - NM_001282649 - c.574_575insGCTCTGGTGGG; p.A192Gfs*100 | P (PVS1, PS2, PM2, PP3) | somatic | 25% [13 - 40%] | - | 0% [0 - 11%] |
| MCD63 | M | FCDI<sup>+</sup> | *SLC35A2* - NM_001282649 - c.C364T;p.Q122X | P (PVS1, PS2, PM2, PP3) | somatic | 1.9% [0.39 - 5.5%] | 2.7% [2.1 - 3.3] | 0% [0 - 3.6%] |
| MCD47 | M | FCDI<sup>+</sup> | *SLC35A2* - NM_001282649 - c.C252A;p.Y84X | P (PVS1, PS2, PM2, PP3) | somatic | 27% [21 - 34%] | 3.8% [3.0 - 4.8%] | - |
| MCD37 | M | FCDI<sup>+</sup> | *SLC35A2* - NM_001032289 - c.G164T;p.R55 | LP (PS2, PM1, PM2, PP3, BP1) | somatic | 54% [39 - 69%] | - | 0% [0 - 5%] |
| MCD67 | F | FCDII | 1qdup/ PTEN - NM_000314 - c.G389A;p.R130Q | P | somatic | -/9% [5.6 - 14%] | -/4.8% [4.4 - 5.2%] | 1.4% [0 - 3.3%] |
| MCD88 | F | FCDII | *DEPDC5* - NM_001369901:c.3938delC: p.A1313Efs*4 | P (PVS1, PS2, PM2, PP3, PP5) | germline | 50% [48 - 53%] | - | 49% [46 - 52%] |
| MCD106 | M | FCDII | *MTOR* - NM_004958 - c.C6644T;p.S2215F | P (PS2, PS3, PM1, PM2, PM5, PP3, PP5, BP1) | somatic | 5.3% [2.9 - 8.9%] | 5.7% [4.6 - 6.9%] | - |
| MCD100 | M | FCDII | *MTOR* - NM_004958 - c.C6644T;p.S2215F | P (PS2, PS3, PM1, PM2, PM5, PP3, PP5, BP1) | somatic | 4.0% [2.3 - 6.6%] | 3.9% [2.8 - 5.4%] | 0% [0 - 1.5%] |
| MCD76 | F | FCDII | *MTOR* - NM_004958 - c.C6644T;p.S2215F | P (PS2, PS3, PM1, PM2, PM5, PP3, PP5, BP1) | somatic | 3.9% [2.7 - 5.4%] | 2.2% [1.8 - 2.7%] | - |
| MCD69 | F | FCDII | *MTOR* - NM_004958 - c.C6644T;p.S2215F | P (PS2, PS3, PM1, PM2, PM5, PP3, PP5, BP1) | somatic | 5.8% [1.2 - 16%] | 3.5% [3.2 - 3.8%] | 0% [0 - 2.6%] |
| MCD80 | F | FCDII | *MTOR* - NM_004958 - c.C6644A;p.S2215Y | P (PS2, PS3, PM1, PM2, PM5, PP3, PP5, BP1) | somatic | 4.7% [2.8 - 7.4%] | 7.4% [3.7 - 7.4%] | 0% [0 - 1%] |
| MCD94 | M | FCDII | *MTOR* - NM_004958 - c.C6644A;p.S2215Y | P (PS2, PS3, PM1, PM2, PM5, PP3, PP5, BP1) | somatic | 7.2% [4.7 - 10%] | - | 0% [0 - 2.4%] |
| MCD99 | M | FCDII | *MTOR* - NM_004958 - c.C6644A;p.S2215Y | P (PS2, PS3, PM1, PM2, PM5, PP3, PP5, BP1) | somatic | 1.9% [0 – 5.4%]*** | 5.2% [4.4 - 6.0%] | 0% [0 – 1.4%] |
| MCD68 | F | FCDII | *MTOR* - NM_004958 - c.C5930G;p.T1977R | LP (PS2, PM2, PM5, PP3, BP1) | somatic | 6.1% [1.2 – 16.8%]*** | 3.1% [2.7 - 2.6%] | 0% [0 – 7.2%] |
| MCD87 | F | FCDII | *MTOR* - NM_004958 - c.C5930A;p.T1977K | P (PS2, PM2, PM5, PP3, PP5, BP1) | somatic | 4.6% [3.3 - 6.4%] | 4.7% [4.1 - 5.3%] | - |
| MCD98 | M | FCDII | *MTOR* - NM_004958 - c.C5930A;p.T1977K | P (PS2, PM2, PM5, PP3, PP5, BP1) | somatic | 3.8 [2.0 - 6.5%] | 2.9% [2.5 - 3.4%] | 0% [0 - 0.9%] |
| MCD104 | M | FCDII | *SCN1A* - NM_001353948 - c.T662C;p.L221P | P (PS2, PM1, PM2, PM5, PP2, PP3, PP5) | germline | 46% [38 - 54%] | - | 42% [35 - 48] |
| MCD70 | F | FCDII | *TSC1* - NM_000368 - c.C298T;p.Q100X | P (PVS1, PS2, PM2, PP3, PP5) | somatic | 4.4% [0.92 - 12] | 4.1% [3.6 - 4.6%] | 1% [0 – 4.4%] |
| MCD74 | F | FCDII | *TSC2* - NM_001318831 - c.1334_1335insCTGCGAC;p.Y448Lfs*11 | P (PVS1, PS2, PM2, PP3) | somatic | 1.7% [1.0 - 2.6%] | - | 0% [0 - 0.6%] |
| MCD115 | F | FCDIII | *PCDH19* - NM_001105243 - c.1958_1959del;p.S653Cfs*66 | P (PVS1, PS2, PM2, PP3, PP5) | somatic | 0.37 [0.12 - 0.86%] | 0.34% [0.31 - 0.37%] | - |
| MCD122 | M | FCDIII | *RANBP2* - NM_006267:c.C1754T;p.T585M | P (PVS1, PS2, PM2, PP5, BP4) | germline | 48% [42 - 53%] | - | - |

Abbreviations: LP=likely pathogenic; P=pathogenic
\*more detailed information about radiographic and pathologic phenotypes are provided in Supplementary Table 1.
\*\*blood sample was not sequenced for this study but this site was assessed in leukocyte-derived DNA clinically
\*\*\*somatic variant not called in our somatic variant calling pipeline

**Table 2.** Clinical phenotypes of novel somatic and germline variants.

| Sample ID | Sex | Candidate diagnosis | VAF | somatic/germline | Seizure types | Preop MRI | Pathology | Localization | Notes |
| --- | --- | --- | --- | --- | --- | --- | --- | --- | --- |
| MCD11 | M | *AKT3* - NM_181690 - c.C863T;p.T288I | 0.42 | somatic | asymmetric myoclonic-tonic spasms | DMEG | mMCD; gliosis | Left parietal-occipital and generalized | on exam: macrocephaly, hypotonia, capillary hemangioma on right back, cutaneous 2-3rd toe syndactyly; strong clinical suspicion for *PIK3CA* segmental overgrowth syndrome |
| MCD59 | M | *ATP10A* - NM_024490 - c.C647A;p.S216X | 0.03 | somatic | focal impaired awareness seizures (staring, perioral cyanosis, +/- right sided stiffening) | FCD NOS | FCD1a | Left temporal sharp prior to surgery 1, Left anterior to midtemporal electroclinical sz |  |
| MCD20 | F | *CASK* - NM_001367721 - c.2317+1G>A | 0.07 | somatic | epileptic spasms | FCD NOS | FCD1a | Right hemisphere onset spasms | No microcephaly or cerebellar atrophy |
| MCD60 | M | *KRAS* - NM_001369786 - c.G35T;p.G12V | 0.2 | somatic | myoclonic seizures→ infantile spasms | FCD NOS vs dysmyelination | FCD NOS | Right temporal lobe |  |
| MCD17 | M | *DEPDC5* - NM_001369901:c.C1675T;p.R559X | ~50% | germline | focal impaired awareness and motor seizures | FCD NOS | Gliosis, FCD1a | Left centroparietal seizures | Hypomelanosis of ito with normal karyotype |
| MCD63 | M | *SLC35A2* - NM_001282649 - | 0.02 | somatic | infantile spasms | FCD NOS | gliosis, FCD1a | Right frontal |  |
|  |  | c.C364T;p.Q122X |  |  |  |  |  |  |  |
| MCD54 | M | *NF1* -<br>NM_000267 -<br>c.1017_1018del;p.<br>S340Cfs*12 | 0.03 | somatic | focal<br>seizures<br>with<br>hyperkinet<br>ic motor<br>automatis<br>ms | NLFE | gliosis<br>(left<br>mesial<br>frontal<br>lobe) | Left frontal | No evidence of<br>neurocutaneous<br>stigmata on<br>examination. No<br>other evaluations to<br>assess for bony<br>abnormalities or<br>other findings of NF1<br>Patient was found on<br>clinical testing to<br>have a germline VUS<br>in NF1 |
| MCD40 | M | *CUL1* -<br>NM_001370660 -<br>c.C187T;p.R63X | 0.072 | somatic | focal<br>impaired<br>awareness<br>seizures | FCD NOS | mMCD | Right temporal | Born at 30 weeks<br>gestation. Cardiac<br>history of secundum<br>atrial septal defect, a<br>patent ductus<br>arteriosus and<br>pulmonary<br>hypertension |
| MCD21 | F | *NIPBL* -<br>NM_015384 -<br>c.7685+1G>A | 0.086 | somatic | focal<br>impaired<br>awareness<br>seizures<br>with<br>secondary<br>generaliza<br>tion | mMCD | mMCD,<br>gliosis | Left frontal<br>electroclinical<br>seizures | No dysmorphic<br>features on exam. |
| MCD88 | F | *DEPDC5* -<br>NM_001369901:c<br>.3938delC;p.A131<br>3Efs*4 | ~0.5 | germline | focal<br>motor<br>seizures,<br>infantile<br>spasms,<br>myoclonic | FCD NOS | FCD IIb | Left<br>hypsarrhythmia,<br>left temporal<br>sharps, left<br>temporal<br>seizures | Developmental delay<br>and right hemianopia,<br>RUE hemiparesis |
| MCD104 | M | *SCN1A* -<br>NM_001353948 - | ~0.5 | germline | Mixed<br>generalize | FCD IIb | FCD IIb | Left frontal<br>electroclinical | Clinically diagnosed<br>as Dravet from, *de* |
|  |  | c.T662C:p.L221P |  |  | d seizures<br>(myoclonic, astatic,<br>GTC with<br>and<br>without<br>fever) |  |  | seizures, left<br>frontal/bifrontal<br>spikes | *novo SCN1A*<br>mutation on GeneDx<br>panel |
| MCD115 | F | *PCDH19* -<br>NM_001105243 -<br>c.1958_1959del;p.<br>S653Cfs*66 | 0.004 | somatic | focal<br>impaired<br>awareness<br>seizures | MTS +/-<br>dysmyelination<br>vs FCD | mMCD* | Right temporal | second seizure<br>episode occurred<br>with strept throat<br>diagnosis |
| MCD122 | M | *RANBP2* -<br>NM_006267:c.C1<br>754T:p.T585M | ~50% | germline | focal<br>impaired<br>awareness<br>seizures | MTS + gliosis<br>(FCD IIIa) | HS | Right temporal | history of possible<br>encephalitis as a<br>young child and was<br>in a coma for 4 weeks<br>(no infectious<br>etiology identified) |

Table abbreviations HS: hippocampal sclerosis; MTS = mesial temporal sclerosis; sz = seizures

One additional HMEG specimen had a copy number neutral somatic 16p uniparental disomy that has been previously reported (Griffin *et al*., 2017). All somatic variants identified in HMEG cases had VAFs greater than 6.7% (average 20%), reflecting a relatively early mutation (Figure 3). MRI for all HMEG cases with a pathogenic somatic variant are provided in Supplementary Figure 1.

**Figure 3.**
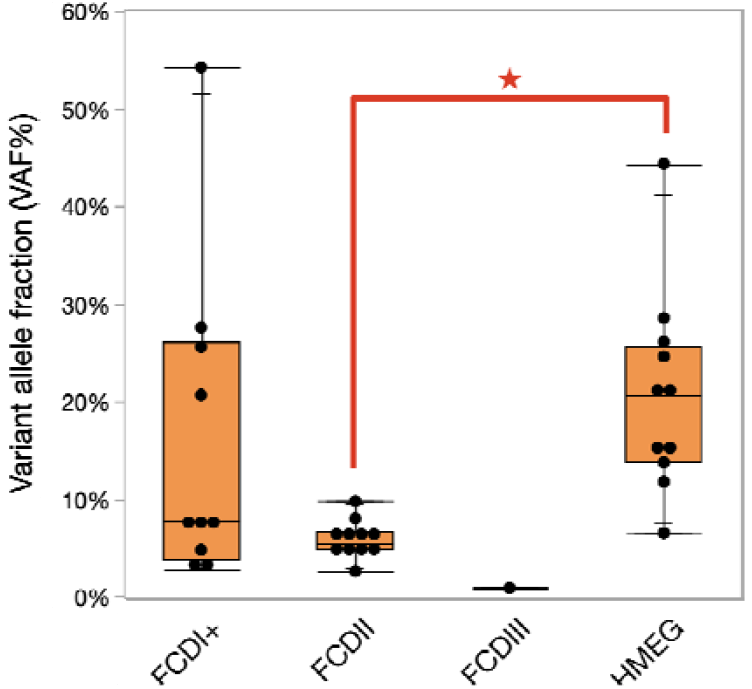
Variant allele fraction (VAF%) as a function of phenotype.

#### FCDII

Among the 44 individuals diagnosed with FCDII, 13 (30%) had explanatory findings. Eight had a pathogenic or likely pathogenic variant detected in the high-quality somatic variant calls in *MTOR* (n=7) or *PTEN* (n=1) (Table 1). The seven individuals with *MTOR* variants harbored either the S2215F (n=3), S2215Y (n=2), or T1977K (n=2) (NM_004958) variants previously reported in individuals with HMEG or isolated FCDII (D’Gama *et al*., 2017). The *PTEN* variant (NM_000314:c.G389A:p.R130Q) was identified in one case with a VAF of 9.2% (Table 1). Interestingly, the individual with FCDIIa harboring the pathogenic *PTEN* variant also had a 1q duplication identified in the CNV analysis (Supplementary Figure 2).

Among the permissive variant call set, we identified another S2215F *MTOR* variant (VAF 5.8%), one somatic *TSC1* variant (NM_000368:c.C298T:p.Q100X, VAF = 4.4%), and one low-level somatic variant in *TSC2* (NM_001318831:c.1334_1335insCTGCGAC:p.Y448Lfs*11, VAF = 1.7%). All were initially not detected because of low coverage at the variant site. Consistent with other reports of individuals with FCDII and somatic *TSC1* and *TSC2* variants (D’Gama *et al*., 2017; Lim *et al*., 2017), neither of these patients met clinical criteria for TSC, even when re-phenotyped in light of these findings.

Two additional cases each with an *MTOR* variant (S2215Y and T1977R) were identified, one with clinical exome sequencing of the resected brain tissue specimen and one with targeted sequencing of *MTOR* that was part of another study (Winawer *et al*., 2018). While both had at least one read supporting the variant in our exome sequence data, neither had reached the threshold required for us to call a somatic variant.

All somatic variants identified in FCDII cases had VAFs less than 10% (average 5%) (Figure 3). Among the ten individuals with pathogenic somatic *MTOR* variants, five had FCDIIa and five had FCDIIb (Supplementary Table 1, Supplementary Figure 3). Both specimens with a somatic *TSC1* or *TSC2* variant had FCDIIb (Supplementary Table 1).

#### FCDI and related phenotypes (FCDI^+^)

We identified one pathogenic variant in *SLC35A2* (VAF 27%) in an individual with mMCD and decreased myelination, a phenotype consistent with that previously reported focal neocortical epilepsy caused by somatic *SLC35A2* variants (Bonduelle *et al*., 2021). In addition, we identified and confirmed disease-causing somatic variants in four additional genes previously implicated in neurodevelopmental disorders. The first of these four cases had a somatic splice site variant in *CASK* (NM_001367721 – c.2317+1G>A) that, when detected as a germline variant, is associated with microcephaly, pontine or cerebellar hypoplasia, and seizures (Najm *et al*., 2008; Piluso *et al*., 2009). This individual exhibited none of the non-neurological phenotypes associated with germline *de novo CASK* variants, consistent with the undetectable levels of the variant allele in the blood (Table 1). Furthermore, this subject showed no evidence of microcephaly or cerebellar atrophy, features commonly observed in individuals with this variant at the time of surgery or 18 months post-surgery (Najm *et al*., 2008) (Figure 4), suggesting that the variant may be localized or enriched in the cortical tissue.

**Figure 4.**
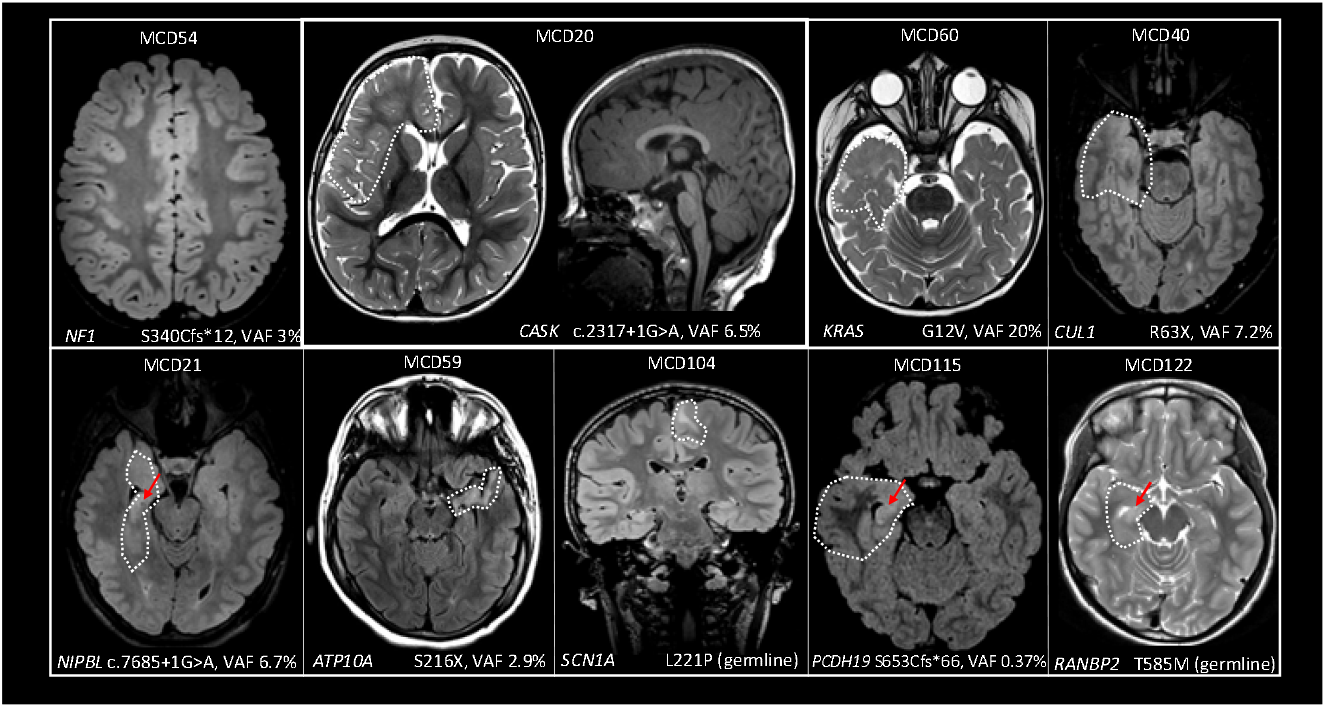
MRI images from MCD cases with a novel variant in *CASK, KRAS, CUL1, NIPBL, ATP10A, SCN1A, PCDH19*, or *RANBP2*.

The second disease-associated variant detected in our FCDI^+^ cohort was a somatic variant in *KRAS* (NM_001369786 – c.G35T;p.G12V), a variant previously implicated in arteriovenous brain malformations (Nikolaev *et al*., 2018), a range of tumors types (Pulciani *et al*., 1982; Kranenburg, 2005), and a recently reported FCD1a case (Pepi *et al*., 2021). Other germline variants in *KRAS* have also been associated with Noonan syndrome and cardio-facio-cutaneous syndrome (Niihori *et al*., 2006; Schubbert *et al*., 2006), both of which can include developmental delay. The patient with the *KRAS* variant had no radiographic evidence of an arteriovenous malformation or tumor, and head circumference was normal. Due to the lack of a blood sample the tissue localization of the post-zygotically acquired *KRAS* variant was not able to be determined, but the patient exhibited none of the non-neurological phenotypes associated with Noonan syndrome or cardio-facio-cutaneous syndrome.

The third likely pathogenic somatic variant identified in FCDI^+^ was a novel somatic splice site variant in *NIPBL*, a gene that gives rise to Cornelia de Lange syndrome I, a disorder associated with highly variable phenotypes including dysmorphism, microcephaly, intellectual disability (ID), and seizures (Tonkin *et al*., 2004). The *NIPBL* variant was not found in leukocyte-derived DNA, and the patient had no clinical features of Cornelia de Lange syndrome I (Figure 4, Table 2).

Finally, the fourth variant identified was a mosaic frameshift deletion in *NF1* (NM_000267: c.1017_1018del:p.S340Cfs*12). Loss-of-function *NF1* variants cause neurofibromatosis 1, a disorder associated with macrocephaly, ID, and seizures (Wallace *et al*., 1991; Valero *et al*., 1994). While leukocyte-derived DNA from this individual with the somatic *NF1* variant was not available for sequencing in this research study, exome sequencing performed clinically on leukocyte-derived DNA did not detect the variant allele despite >500-fold coverage (Table 1). This individual had no clinical features suggestive of neurofibromatosis 1 (Figure 4, Table 2). However, in addition to the somatic variant identified in this study, a *NF1* germline variant of unknown significance (VUS; NM_000267:c.G4122T:p.Q1374H) was identified during clinical exome sequencing testing in the individual with a somatic *NF1* variant. “Second hits” in *NF1* have been previously identified in benign neurofibromas (Serra *et al*., 1997), although not specifically to our knowledge within an epileptic zone of an individual with focal epilepsy.

One low-level (VAF 0.03) VUS was identified and confirmed in *ATP10A* (NM_024490:c.C647A:p.S216X) (Table 1). This variant appeared in two separate exome sequencing runs of two specimens from the same individual. No other sample was sequenced multiple times and no other somatic variant identified in this case was found in both sequencing runs. *ATP10A* encodes a phospholipid-transporting ATPase and is not a known disease gene. At a population level, loss-of-function variants in this gene were identified slightly less frequently than expected in the gnomAD database (loeuf score of 0.43), which may indicate that this class of variants in *ATP10A* may be under negative selection (Karczewski *et al*., 2020). The specimen from this individual had pathology suggesting FCDIa with disrupted cytoarchitecture and neuronal heterotopia (Table 2).

Since several of the previously reported and confirmed *SLC35A2* variants identified from the same sequencing data were not called in high-quality variant calls, we also evaluated a more permissive set of calls (see Methods) whereby we relaxed the read depth requirements and variant allele fraction thresholds and excluded only variant calls that are likely due to sequencing or alignment errors. Given the likely high number of false positives we expect in the permissive call set, we limited our analyses only to known pathogenic variants or variants in known MCD genes. Using this approach, we identified three additional somatic *SLC35A2* variants in three different individuals in the FCDI^+^ cohort, including one novel low-level somatic variant in *SLC35A2* (NM_005660:c.C547T; VAF=0.02) and two somatic *SLC35A2* variants (NM_005660:c.757_758insGCTCTGGTGGG:p.A253Gfs*100 and c.G164T:p.R55L) that were previously identified and reported on in another publication (Winawer *et al*., 2018). All had not been initially identified because of low coverage at the variant site, lowering confidence that the somatic variant was truly present. One of the cases with a somatic *SLC35A2* variant had mMCD, whereas the others exhibited very subtle cortical dyslamination though not meeting formal diagnostic criteria for FCDIa or Ib (Supplementary Table 1).

Given that we had exome sequence data from a matched leukocyte-derived DNA sample for a fraction of the cohort, we also reviewed somatic variant calls from the matched sample to ensure that no somatic variants were identified only in a paired brain-blood analysis. None were identified in the FCDI^+^ cohort.

Only one high-quality somatic CNV was identified in the FCDI^+^ cohort, a chromosome 1q duplication (Supplementary Figure 2). Chromosome 1q duplications have been reported in individuals with hemimegalencephaly, (Poduri *et al*., 2012) and unilateral PMG with dysmorphic neurons (Kobow *et al*., 2020). This individual was diagnosed with FCDNOS with heterotopic white matter neurons and pancortical eosinophilic astrocytic inclusions, a pattern described in detail [see case 2) in (Fischer *et al*., 2020)].

#### FCDIII

In FCDIII, we identified only one somatic variant among the 16 cases evaluated. This individual has FCD plus radiographic evidence of mesial temporal sclerosis (Figure 4) and had a very low level mosaic (VAF = 0.4%) loss-of-function *PCDH19* variant (NM_001105243: c.1958_1959del:p.S653Cfs*66) identified in the cortical specimen from that patient that was identified in the permissive variant call set and confirmed with deep amplicon sequencing.

#### Germline variant analysis

We identified a total of four likely germline diagnoses (Table 1). Two of the pathogenic germline variants were found in *DEPDC5*, NM_001369901:c.C1675T:p.R559X and c.3938delC:p.A1313Efs*4 both with probable FCDIa.(Scheffer *et al*., 2014; Baulac *et al*., 2015). Somatic second hits were not identified in either of the germline *DEPDC5* individuals, although we cannot rule out that they were undetected due to low VAF or technical limitations. The third individual had a known pathogenic germline *SCN1A* variant (NM_001353948 – c.T662C:p.L221P) and had been diagnosed radiographically and by pathology with FCDIIb. The *SCN1A* variant was also found to be *de novo* with clinical sequencing after inclusion in this study. While not previously associated with FCDII, the *SCN1A* variant was deemed by a diagnostic laboratory to be likely contributing to the individual’s epilepsy. Finally a fourth variant in *RANBP2* (NM_006267:c.C1754T:p.T585M) was identified in an individual with FCDIII, specifically FCD with pathology consistent with hippocampal sclerosis (Blumcke *et al*., 2013). This *RANBP2* variant is associated with acute infection-induced encephalopathy (Neilson *et al*., 2009) and indeed, on further review of the clinical records, the patient had a history of a coma due to presumed encephalitis in early life; an infectious etiology was never identified. The patient developed intractable neocortical seizures that resulted in surgical resection before the age of 10.

### Novel gene search

After completing a targeted evaluation for variants in known genes, we next sought to evaluate the high-quality call set for candidate somatic variants that may contribute to the etiological landscape of the MCDs. The majority of pathogenic variants in dominant acting genes involved in rare neurodevelopmental conditions, such as epilepsy and brain malformations, are ultra-rare in the population, putatively functional, and are often found in intolerant or constrained genes, i.e. genes that tend to be depleted of functional variants, specifically loss-of-function variants, in the general population (Epi4K Consortium and Epilepsy Phenome/Genome Project, 2013; Petrovski *et al*., 2013; Samocha *et al*., 2014). Given this observation, we first evaluated only those variants that are rare (gnomAD exome MAF < 1 × 10^−6^), functional [splice-site, nonsense, missense excluded Polyphen (HumVar) benign, frameshift, and nonframeshift indel], and located in a constrained gene, or a rare loss-of-function variant (nonsense, splicing, or frame-shift indel) in a gene with a pLI> 0.9 (Samocha *et al*., 2014). Across all 123 cases, there were 49 variants meeting these criteria. There were only two genes that harbored more than one high-quality somatic variant call confirmed with independent genotyping in different individuals, including *MTOR* (n=10) and *PIK3CA* (n=5). Each of these *MTOR* and *PIK3CA* variants had been identified in the diagnostic analyses. Among the genes with single somatic variants identified, five had been identified in the diagnostic analysis above (*AKT3, CASK, NF1, NIPBL*, and *PTEN*). Among the 31 qualifying variants remaining, all except one were not felt to be strong candidates either because they appeared to be an artifact with visual inspection, were found to be more common in other control datasets, did not confirm with secondary genotyping, or were in genes unlikely to contribute to disease. The one additional candidate identified and confirmed in this analysis was a somatic loss-of-function variant in *CUL1* (NM_001370660:c.C187T:p.R63X, VAF = 7.2%). *CUL1* encodes a protein comprising the ubiquitin ligase complex that serves to ubiquitinate proteins involved in cell cycle progression. *CUL1* has not to date been implicated in human disease, but it is embryonic lethal when knocked out in mice (Wang *et al*., 1999).

### Aggregate analyses

#### Enrichment of known pathogenic somatic variants in MCD

Given the lack of knowledge of the somatic genetic landscape in brain tissue derived DNA from healthy individuals, we next sought to compare the rate of low level pathogenic somatic variants in MCD cases to that of controls. The control cohort used for this analysis consisted of exome and whole genome sequence data from autopsy-collected brain tissue specimens from 63 individuals without neurological disorders made available for biomedical research through the North American Brain Expression Consortium and the Brain Somatic Mosaicism Network. To ensure a fair comparison between cases and controls, we identified a list of 15,622 out of 18,078 CCDS genes where that were sequenced to approximately equal depths and extents in cases and controls (Supplementary Table 6, see Methods). Evaluating only these 15,622 genes we performed the somatic diagnostic analysis (see Methods) initially applied to our MCD cases in the control cohort. Because DNA was not available to us to confirm variant calls in controls we did not consider variant validation status in our analysis and only relied on the bioinformatically-defined filtering criteria that could be applied consistently across both cohorts. Within the coverage normalized gene-set (15,622 genes), we found no candidate somatic genetic diagnoses in controls, whereas in the FCDI^+^, FCDII, and HMEG cohorts we identified 5/46 [11%, 2-sided Fisher’s exact test (FET) *P* = 0.017], 7/39 (18%, *P*, = 8 × 10^−4^, and 10/15 (67%, *P* = 2.4 × 10^−9^), respectively. Not unexpectedly, the enrichment of somatic genetic diagnoses in the FCDII and HMEG cohorts was driven by somatic variants in *PIK3CA, AKT3*, and *MTOR*. The enrichment in FCDI^+^ was driven by the singleton variants discussed in the diagnostic analysis in *NF1, NIPBL, KRAS, SLC35A2*, and *CASK* and suggest that these variants are contributing to the somatic genetic risk of FCDI^+^. Even after removing the one variant in the only known FCDI^+^ associated gene, *SLC35A2*, there were still more than expected somatic genetic variants observed in the FCDI^+^ cohort compared to controls (FET *P* = 0.028), also suggesting the presence of novel pathogenic genetic variants present in our dataset.

#### Pathway enrichment analyses

Finally, we performed a pathway analysis to evaluate if high-quality somatic variants were enriched in biologically informed gene sets in any of the MCD cohorts. Across 36 KEGG-defined neurological pathways, we observed no enrichment of somatic variants in cases compared to controls in the FCDI^+^ and FCDIII cohorts. Not unexpectedly, within FCDII and HMEG somatic variants in the PI3K-AKT3-mTOR signaling pathway genes were observed more than expected (Figure 5). The Ras signaling pathway genes had more than expected somatic variants in HMEG cases, although the signal was driven by variants in *AKT3* and *PIK3CA* (Figure 5).

**Figure 5.**
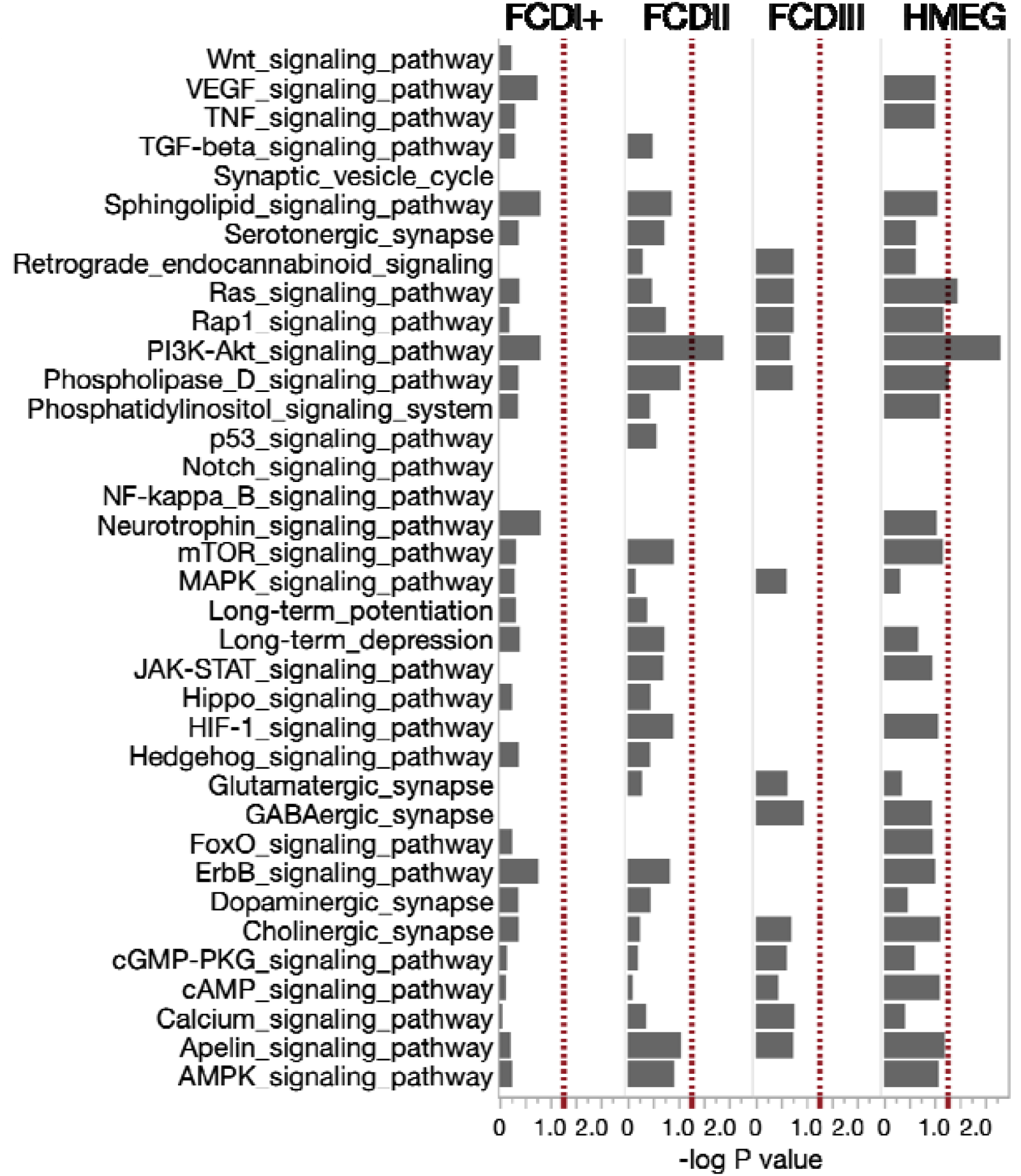
DNENRICH analysis. Enrichment of somatic variants in KEGG-defined pathways in cases compared to controls.

## Discussion

The role of post-zygotically acquired somatic variants that arise during fetal brain development in epilepsy-associated MCD has been increasingly recognized over the past several years. Prior reports of somatic variants have focused largely on deep sequencing of genes known to be involved in specific types of focal cortical malformations. Here we report the results of the first and largest genome-wide investigation for post-zygotically acquired somatic genetic variants in the resected brain tissue of 123 patients with epilepsy. Based on rigorous phenotypic assessment of their MRI and neuropathological findings, we classified the lesions into four clinically distinct forms of MCD—FCD^+^, FCDII, FCDIII, and HMEG—which allowed for a novel comparison of the genetic profiles of different FCD types.

Similar to what we and others have previously reported, we identified a high rate of pathogenic or likely pathogenic genetic findings in HMEG (75%) as well as the commonly encountered and radiographically and pathologically well-defined FCDII (30%) (Figure 6). All pathogenic and likely pathogenic variants that we identified in HMEG and FCDII cases were in genes comprising the PI3K-AKT-mTOR signaling pathway, which is not surprising given the shared pathological features between these malformations. The variants in *AKT3, DEPDC5, MTOR, PI3KCA, PTEN, RHEB, TSC1*, and *TSC2* in HMEG and FCDII reinforce the pivotal role of mTOR hyperactivation as a shared mechanism underlying these select MCD subtypes (Lee *et al*., 2012; Poduri *et al*., 2012; D’Gama *et al*., 2015; Nakashima *et al*., 2015; Lim *et al*., 2017; Baldassari *et al*., 2019; Salinas *et al*., 2019; Zhao *et al*., 2019), substantiating their characterization as ‘mTORopathies’ (Crino, 2015).

**Figure 6.**
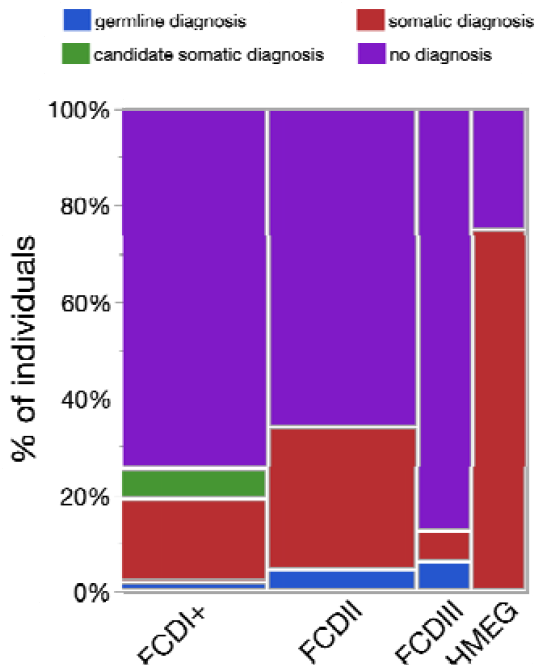
Rate of genetic diagnoses across phenotypic categories. Percent of individuals with a germline genetic diagnosis (blue), a somatic genetic diagnosis (red), a candidate somatic genetic diagnosis (green), or no genetic diagnosis identified (purple). The size (width) of the bar corresponds with size of the cohort.

While somatic gene discovery in FCDII has progressed rapidly in the past few years, identifying specific causes for FCDI has been more challenging. As we begin to identify somatic variants as the cause of FCDI, it is clear multiple distinct genes associated with brain development lead to FCDI. This genetic heterogeneity for FCDI is in sharp contrast to FCDII, for which virtually all associated gene variants have been linked to the PI3K-AKT-mTOR signaling pathway. Despite the contrasting genetics between FCDI and FCDII, we did identify a single individual with FCDNOS (FCD1^+^ category) with a somatic chromosome 1q amplification, a CNV that has been reported previously in HMEG (Poduri *et al*., 2012) and two individuals with a *DEPDC5* variant, one with FCD1a and one with FCDII. Our findings could suggest a possible link between these seemingly distinct clinical entities, however other confounding factors may make interpretation of the phenotypes of these two genes more challenging. For 1q ampliciation, the variable phenotype may be caused by excess expression of *AKT3*, although it may equally likely be due to combined effects of triplosensitivity of a large number of genes residing on the q arm of chromosome 1 and/or variable cell-type specific mutation burdens. For the *DEPDC5* individuals, the variability may be explained by the presence or absence of a second hit, as has been suggested previously (Lee *et al*., 2019). While no second hit was identified in this study it does not mean that they were present and simply undetected at the sequencing depth achieved in these cases. Despite the challenges identifying genes in FCDI and related phenotypes, we note that the rate of genetic diagnosis in FCDI^+^ approaches that of FCDII when considering the novel findings identified through this work (Figure 6).

Consistent with prior studies of lesional and non-lesional focal neocortical epilepsy, we identified somatic variants in *SLC35A2* in FCDI^+^ cases in this cohort. Three of the four individuals with *SLC35A2* had pathological features consistent with MOGHE. Pathological assessments of these cases at the time of ascertainment did not specifically reference MOGHE since this was not well recognized as a distinct pathological entity and still is not a formal ILAE classification term. Additional quantitative studies will likely be needed to develop more definitive diagnostic guidelines that take into account developmental and region-specific oligodendrocyte development (Chang *et al*., 2000; Zerlin *et al*., 2004). Interestingly, we also identified somatic variants in *KRAS, NF1, NIBPL*, and *CASK*, genes previously associated with neurodevelopmental disorders but in the presence of, *de novo* germline variants. All of the somatic variants identified were either identical to the previously reported pathogenic germline *de novo* variants (*CASK* and *KRAS*) or were predicted to result in the same functional effect on the protein (*NF1, NIPBL*, and *SLC35A2*). The phenotypes associated with each of these gene variants in our MCD patients were restricted to brain-associated phenotypes, not surprisingly, since the variants were mosaic and enriched in brain tissue in all cases where a blood sample was available (Table 1). For example, the patient with the somatic *NIPBL* variant lacked limb abnormalities and facial dysmorphisms, but shared seizures, brain malformation, and developmental delay associated with germline *de novo* variants in *NIPBL* in patients with Cornelia de Lange syndrome (Tonkin *et al*., 2004). These examples provide additional evidence, similar to that seen with *SLC35A2*, that at least some genes with germline variants previously implicated in neurodevelopmental disorders but not focal brain malformations, can nonetheless cause epilepsy-associated focal MCD when they occur as somatic variants.

*NF1, NIPBL*, and *KRAS* play known roles in normal neuronal development. For example, approximately 50% of autopsied brains from neurofibromatosis patients show disordered cortical architecture with random orientation of neurons, focal heterotopic neurons, proliferation of glial cells to form well-defined gliofibrillary nodules, and hyperplastic gliosis (Rosman and Pearce, 1967). In addition, *NF1* knockout mice have reduced cerebral cortical thickness and increased cell packing density (Zhu *et al*., 2001). Mouse embryonic brains *in utero* electroporated with *Nipbl*-targeting shRNAs at embryonic day 14.5 exhibit defects in neuronal migration including significant accumulation of transfected neurons in the intermediate zone and a reduction of cell numbers in the cortical plate (van den Berg *et al*., 2017). Interestingly, the somatic *KRAS* variant detected in FCDI has been specifically implicated in arteriovenous malformations when localized to brain endothelial cells (Nikolaev *et al*., 2018), and more recently, reported in a single case with FCDIa (Pepi *et al*., 2021). The former raises questions about the impact of cell-type specific variant burden in dictating phenotype and highlights an area where additional research is needed.

FCDIII is pathophysiologically more complex than FCDI and II, since it is characterized by FCDI^+^ associated with an additional neuropathological finding i.e. vascular malformation, glioneuronal tumor, or hippocampal sclerosis (Blumcke *et al*., 2011). Our gene discovery in FCDIII is completely novel since no study to date has identified a genetic association with this MCD subtype. In our study, we identified a single very low level (VAF ∼0.4%) somatic *PCDH19* variant in the temporal cortical sample a female individual classified as FCDIII based on the presence of a mild FCD in the presence of mesial temporal sclerosis. This patient had seizure onset before 5 years of age and neurobehavioral difficulties; she had normal overall cognitive ability but relative weakness in verbal conceptual skills, which is milder than what is typically seen in germline *PCDH19*-related epilepsy in females (Smith *et al*., 2018). We also identified a case of *RANBP2-*associated encephalopathy (Neilson *et al*., 2009) that was linked to an undefined infectious illness, and later developed into focal epilepsy with mesial temporal sclerosis. Since mesial temporal sclerosis may represent the secondary effects of repetitive or prolonged seizures, it is possible that it represents a secondary finding that can influence the classification of the FCD from FCDI (what they would have been called without the mesial temporal sclerosis) to FCDIII. If that is the case, we would expect overlap between the genetics of FCDI and FCDIII to emerge over time.

Our study also suggests that while low-level pathogenic somatic variants can be detected, a considerable fraction of FCDI, FCDII, and FCDIII remain genetically unexplained despite high-depth exome sequencing. In our study, the variant allele fraction (VAF) averaged around 20% for HMEG, compared to less than 5% in FCDII which are much smaller brain malformations (Figure 3). Even within individuals specifically with pathogenic *MTOR* variants, larger lesions were observed with higher VAFs (Supplementary Figure 3). This observation aligns with the previously reported relationship between lesion size and balloon cell burden with VAF in brain malformations associated with mTOR-associated pathologies (D’Gama *et al*., 2017; Baldassari *et al*., 2019). In addition, we observed two instances in which a pathogenic somatic *MTOR* variant was initially missed because of a combination of the low level of the variants and insufficient depth of sequencing at the locus to detect the mutation, one case of an extremely low-level pathogenic somatic *PCDH19* variant in an individual with FCDIII, and multiple somatic variant calls in FCDI^+^ with VAFs less than 5%. These data collectively suggest that even with high-depth exome sequencing, pathogenic somatic variants may be missed that may be captured with even higher depth sequencing, single cell sequencing, or more advanced sequencing and variant calling approaches to allow for more accurate calling of low-level somatic variants. Such low-level variants may explain at least a subset of the ∼67% of genetically-unexplained individuals in our study.

The translational potential for our findings includes the prospect of precise genetic diagnosis for patients with focal cortical malformations which will require clinical laboratories to not only be able to detect but also to be able to confidently curate and validate somatic variants, including those present at very low allele fraction, which is not current practice. Further, since MCDs typically present with refractory epilepsy, there is potential for not only diagnostic precision but also therapies predicated on the genetic abnormality that not only caused the cortical malformation but that might also be responsible for establishment and maintenance of the epileptic network. We are closest to this translational goal with the HMEG and FCDII categories of MCD. Since non-mTOR-related genetic etiologies for HMEG and FCDII are rare but not completely absent, e.g. associated with the autosomal recessive *CNTNAP2* syndrome (Strauss *et al*., 2006), and a portion of HMEG and FCDII remain currently unexplained, we cannot say that these diagnoses are exclusively mTORopathies. However, as deeper sequencing techniques and interrogation of non-coding sequence become more sensitive to detect low-level mosaicism of PI3K-AKT-mTOR pathway gene variants, we anticipates further solidification of the HMEG/FCDII malformations as a spectrum of mTORopathies, which will have the potential for therapeutic intervention with mTOR inhibitors as has been implemented in TSC.

In conclusion, our study has shed important new light on the somatic genetic bases of MCD, most notably emerging causes for FCDI and related phenotypes in the form of somatic variants in a diverse set of genes involved in neuronal development. While FCDII and HMEG have been well-characterized by MRI, pathology, and now increasingly genetic causes involving the mTOR pathway, FCDI and FCDII are more heterogeneous disorders, and we are only at the beginning of achieving diagnostic precision in this research arena, which also will require translation to the clinical evaluation of patients with focal epilepsy. Our findings further support the incorporation of genetic findings, in addition to the currently used pathological classification, into stratifying diverse forms of MCD and motivates even larger scale studies to further define the somatic genetic landscapes of MCD.

## Data Availability

We are in the process of making the data available through dbgap. In the meantime we will make separate direct arrangements for reasonable requests while the data gets submitted to dbgap.

## Abbreviations

CCDS: consensus coding sequence
DMEG: dysplastic megalencephaly
FCD: focal cortical dysplasia
FCDNOS: focal cortical dysplasia not otherwise specified;, HMEG = hemimegalencephaly
ID: intellectual disability
indel: insertion or deletion variant
LP: likely pathogenic
MCD: malformations of cortical development
MAF: minor allele frequency
mMCD: mild malformation of cortical development
NLFE: nonlesional focal epilepsy
P: pathogenic
SNV: single nucleotide variant
CNV: copy number variant
VAF: variant allele fraction
VUS: variant of unknown significance

## Acknowledgements

The authors kindly thank all the study participants and their families for their willingness to contribute samples and clinical data to this work.

The authors also thank the Bioinformatics and Analytics Resarch Collaborative, and in particular Dr. Hemant Kelkar and Halina Krzystek for their guidance developing and implementing the bioinformatics pipeline used in this study. We also thank Mr. Corey Bailey in the Duke Pathology Department for his extensive assistance with locating and organizing archived slides for review of clinical cases.

Data and/or research tools used in the preparation of this manuscript were obtained from the National Institute of Mental Health (NIMH) Data Archive (NDA). NDA is a collaborative informatics system created by the National Institutes of Health to provide a national resource to support and accelerate research in mental health. Dataset identifier(s): 2962. This manuscript reflects the views of the authors and may not reflect the opinions or views of the NIH or of the Submitters submitting original data to NDA.

## Supplementary material

### Supplmentary Figures

**Supplementary Figure 1. MRI images from HMEG cases with a somatic chr16 UPD, *AKT3, MTOR*, and *PIK3CA* variants**.

**Supplementary Figure 2. Chromosome 1q duplications**. Log-fold change and variant allele frequency (CNVradar) genome-wide (A and C) and chromosome 1 (B and D) for MCD67 (FCDII; A and B) and MCD39 (FCDI+; C and D).

**Supplementary Figure 3. MRI images from MCD cases with a somatic MTOR variants. Images are provided in order of lowest to highest VAF (A ->M)**.

### Supplmentary Tables

**Supplementary table 1**. Radiographic and pathological phenotypes of MCD cases.

**Supplementary table 2**. Control cohort

**Supplementary table 3**. Malformation of cortical development cohort

**Supplementary table 4**. Databases used for annotation

**Supplementary table 5**. Gene lists used in the analyses

**Supplementary table 6**. Coverage normalization across MCD cases and controls

**Supplementary table 7**. High quality somatic calls

