## Supplementary figures and images for "Somatic mutation involving diverse genes leads to a spectrum of focal cortical malformations"

### supplementary figures

Supplementary Figure 1

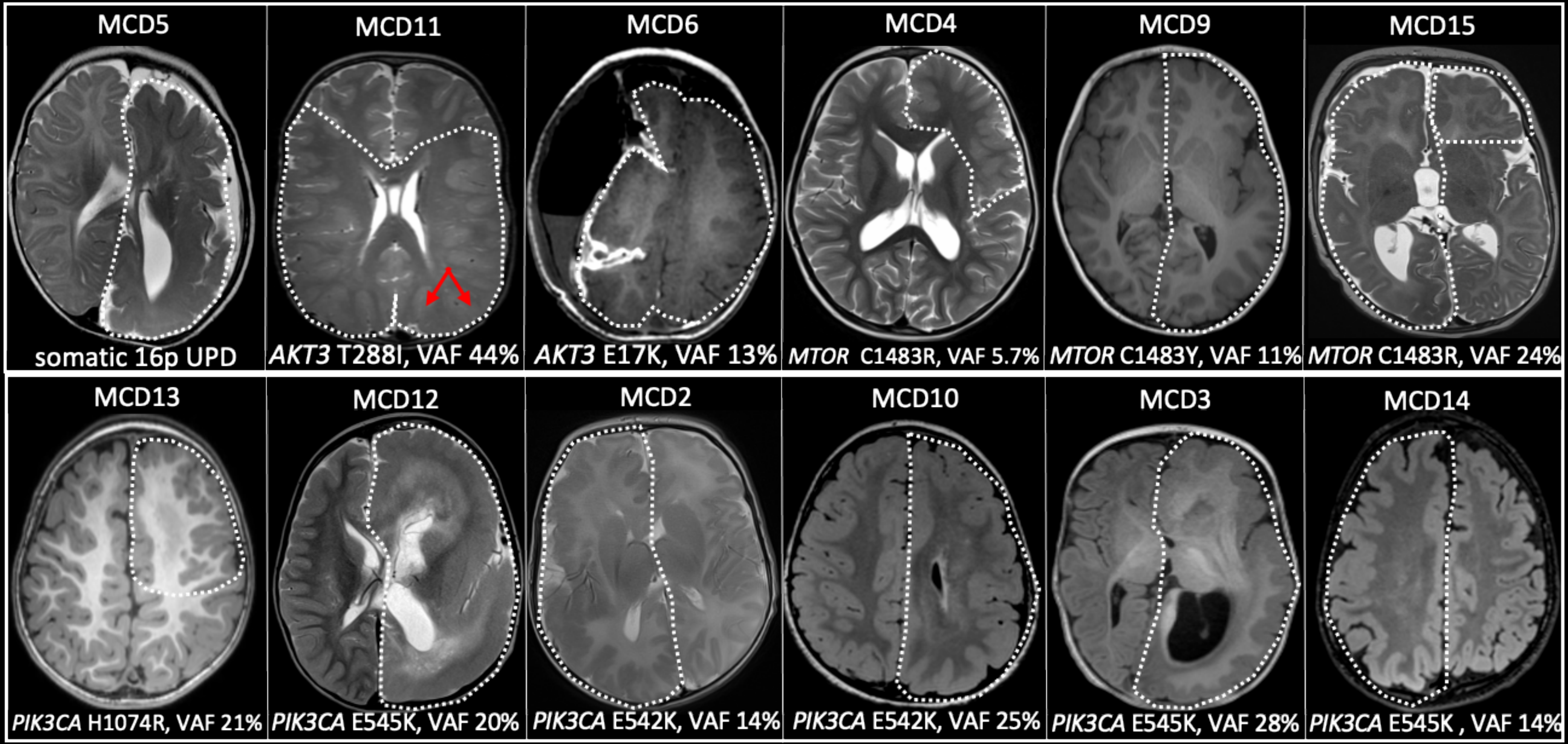

Supplementary Figure 2

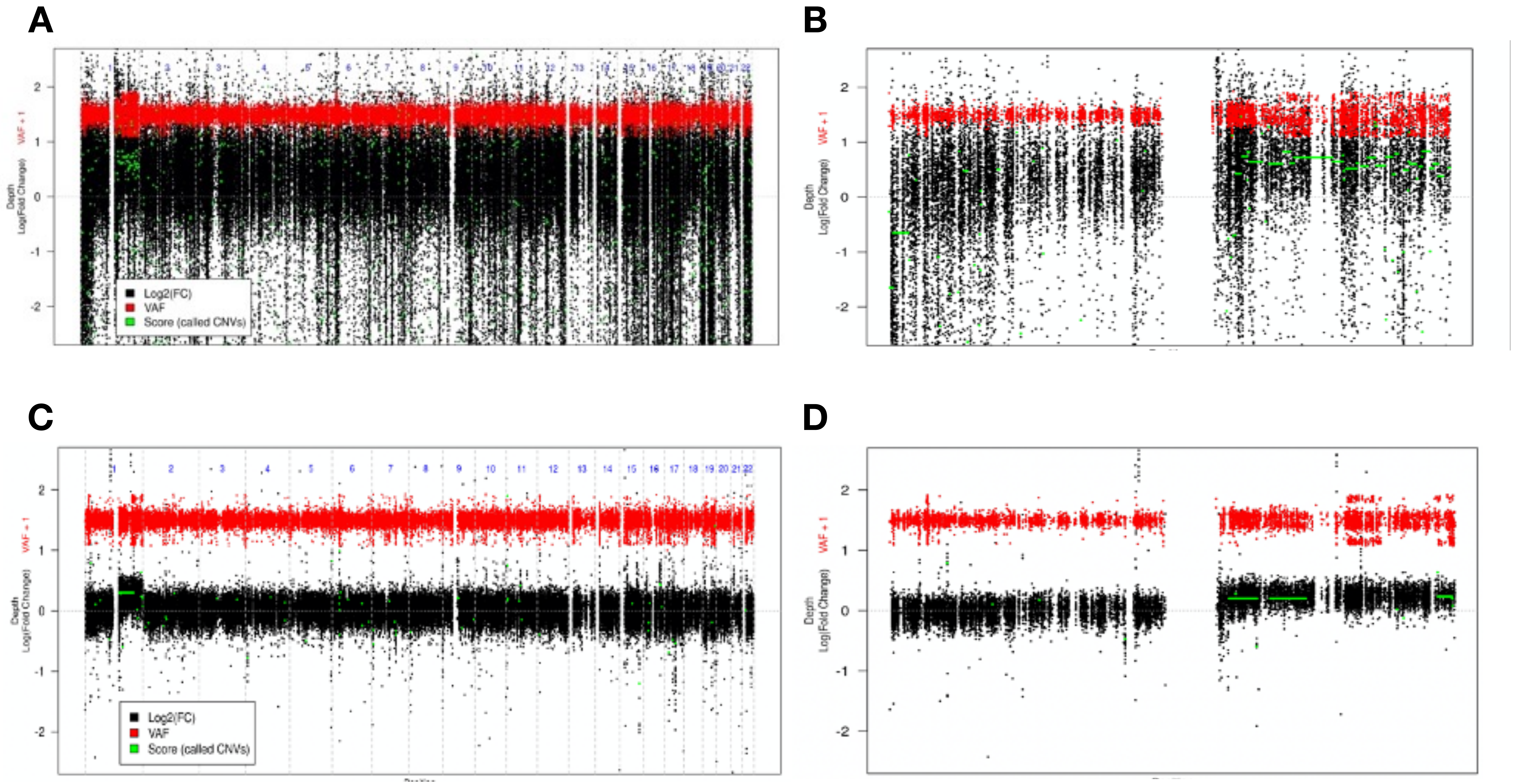

Supplementary Figure 3

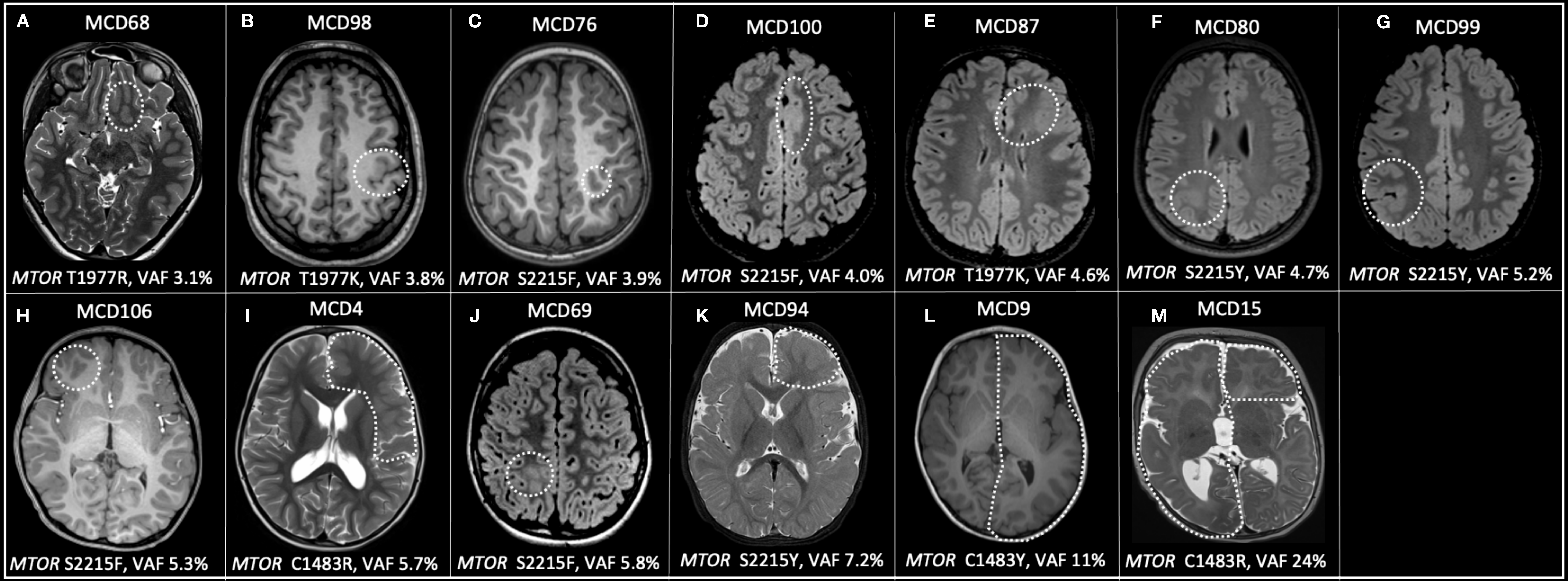
